# Clinical Outcomes in Hospitalized Patients with COVID-19 on Therapeutic Anticoagulants

**DOI:** 10.1101/2020.08.22.20179911

**Authors:** Niti G. Patel, Ajay Bhasin, Joseph M. Feinglass, Steven M. Belknap, Michael P. Angarone, Elaine R. Cohen, Jeffrey H. Barsuk

**Author notes:** **Correspondence**: Niti Patel 251 E. Huron Street, Feinberg 16-738, Chicago, Illinois 60611. **Funding/Support:** None.

## Abstract

**Background:** COVID-19 is associated with hypercoagulability and an increased incidence of thrombosis. We evaluated the clinical outcomes of adults hospitalized with COVID-19 who either continued therapeutic anticoagulants previously prescribed or who were newly started on anticoagulants during hospitalization.

**Methods:** We performed an observational study of adult inpatients’ with COVID-19 at 10 hospitals affiliated with Northwestern Medicine in the Chicagoland area from March 9 to June 26, 2020. We evaluated clinical outcomes of subjects with COVID-19 who were continued on their outpatient therapeutic anticoagulation during hospitalization and those who were newly started on these medications compared to those who were on prophylactic doses of these medications based on the World Health Organization (WHO) Ordinal Scale for Clinical Improvement. The primary outcome was overall death while secondary outcomes were critical illness (WHO score ≥5), need for mechanical ventilation, and death among those subjects who first had critical illness adjusted for age, sex, race, body mass index (BMI), Charlson score, glucose on admission, and use of antiplatelet agents.

**Results:** 1,716 subjects with COVID-19 were included in the analysis. 171 subjects (10.0%) were continued on their outpatient therapeutic anticoagulation and 201(11.7%) were started on new therapeutic anticoagulation during hospitalization. In subjects continued on home therapeutic anticoagulation, there were no differences in overall death, critical illness, mechanical ventilation, or death among subjects with critical illness compared to subjects on prophylactic anticoagulation. Subjects receiving new therapeutic anticoagulation for COVID-19 were more likely to die (OR 5.93; 95% CI 3.71-9.47), have critical illness (OR 14.51; 95% CI 7.43-28.31), need mechanical ventilation (OR 11.22; 95% CI 6.67-18.86), and die after first having critical illness. (OR 5.51; 95% CI 2.80 −10.87).

**Conclusions:** Continuation of outpatient prescribed anticoagulant was not associated with improved clinical outcomes. Therapeutic anticoagulation for COVID-19 in absence of other indications was associated with worse clinical outcomes.

## INTRODUCTION

Adults hospitalized with COVID-19 are at increased risk of thromboembolic disease (1–8). This has been attributed to COVID-19-associated blood hyperviscosity (9) and vascular endotheliïtis (10-12), which suggests that therapeutic strategies directed solely at the coagulation cascade, may be inadequate. A study of an ICU using routine duplex ultrasonography of the lower extremity in critically-ill COVID-19 patients found a high incidence of venous thromboembolism despite therapeutic anticoagulation (13).

There are no randomized controlled trials and few observational cohort studies addressing the potential benefits and harms of anticoagulating hospitalized patients with COVID-19. Paranjpe et al. reported (14) that therapeutic anticoagulation treatment reduced COVID-19-associated mortality among patients on mechanical ventilation, while Tang et al. reported (15) that anticoagulation treatment reduced COVID-19-associated mortality among patients with sepsis-induced coagulopathy. However, both of these prior observational studies failed to account for survivorship bias; and the analysis by Tang et al. lacked generalizability as three-quarters of screened patients were excluded from their analysis (16,17). Despite the paucity of available evidence, a recent CHEST Guideline and Expert Panel Report recommended that hospitalized COVID-19 patients who are acutely-ill or critically-ill and without a contraindication to anticoagulants be prescribed anticoagulant thromboprophylaxis with low-molecular-weight heparin (LMWH) or fondaparinux (18).

In this study, we evaluated the outcomes of adults hospitalized with COVID-19 who either continued therapeutic anticoagulants previously prescribed or who were newly started on anticoagulants during hospitalization. We hypothesized that patients whose previously prescribed therapeutic anticoagulants were continued during hospitalization would have better outcomes than those who were on prophylactic anticoagulation while hospitalized.

## METHODS

We performed an observational cohort study of hospitalized adults with COVID-19 from ten hospitals affiliated with Northwestern Medicine between March 9 and June 26, 2020. Northwestern Medicine is an integrated academic health system in Chicagoland. We evaluated clinical outcomes among subjects who continued their home therapeutic anticoagulation regimen and those who were newly started on anticoagulants to those who were started on prophylactic anticoagulation during their COVID-19 hospitalization. Clinical outcomes were classified according to the World Health Organization (WHO) Ordinal Scale for Clinical Improvement (Table 1) (19). The Northwestern University Institutional Review Board approved this study (Approval Reference STU00212532).

**Table 1:** World Health Organization (WHO) Ordinal Scale for Clinical Improvement.

| Patient state | Descriptor | Score |
| --- | --- | --- |
| Uninfected | No clinical or virological evidence of infection | 0 |
| Ambulatory | No limitation of activities | 1 |
|  | Limitation of activities | 2 |
| Hospitalized mild disease | Hospitalized, no oxygen therapy | 3 |
|  | Oxygen by mask or nasal prongs | 4 |
| Hospitalized severe disease | Non-invasive ventilation or high-flow oxygen | 5 |
|  | Intubation and mechanical ventilation | 6 |
|  | Ventilation + additional organ support-<br>vasopressors, Renal Replacement Therapy,<br>Extracorporeal membrane oxygenation | 7 |
| Dead | Death | 8 |

### Procedure

Data was extracted from the Northwestern University Enterprise Data Warehouse (NUEDW), an integrated database derived from the clinical data of all patients at our health system’s 10 hospitals, each of which use electronic medical records exclusively. The study cohort comprised adults (age ≥18 years old) hospitalized with COVID-19 (ICD-10 code) who were either discharged or died during the study period. We evaluated subjects who had been prescribed therapeutic anticoagulation prior to hospitalization and who then continued therapeutic anticoagulation during hospitalization. We also evaluated subjects who had not been prescribed therapeutic anticoagulation prior to hospitalization and who then began therapeutic or prophylactic anticoagulation during hospitalization. The anticoagulants specified in the database included those used at Northwestern Medicine: apixaban, argatroban, bivalirudin, dabigatran, dalteparin, edoxaban, enoxaparin, fondaparinux, heparin, rivaroxaban, and warfarin. One author (NGP) manually reviewed the records for the index hospitalization for each subject who received at least one dose of therapeutic anticoagulation to determine the indication for anticoagulation and whether therapeutic anticoagulation had been prescribed prior to admission or was newly started while hospitalized. We categorized subjects into the following six groups: continued home therapeutic anticoagulation, started therapeutic anticoagulation for new venous thromboembolism (VTE) during hospitalization, started therapeutic anticoagulation for new atrial fibrillation during hospitalization, started therapeutic anticoagulation for concern of COVID-19 induced vasculopathy (no known other indication) during hospitalization, prophylactic dose anticoagulation during hospitalization, or no anticoagulation during hospitalization. We also collected information on age, sex, race, body mass index (BMI), Charlson score, admission blood glucose readings, use of antiplatelet agent (aspirin, clopidogrel, prasugrel, or ticagrelor), ICD-10 codes associated with admission, and outcomes from the WHO Ordinal Scale (Table 2). Another author (JHB) performed manual chart review on missing admission glucose levels because the original NUEDW query did not return an admission plasma glucose for a small number of subjects. Admission blood glucose was assessed because hyperglycemia is an independent predictor of mortality in patients with COVID-19, regardless of diabetes diagnosis (20–22). For subjects with multiple admissions, the hospitalization with the worst WHO outcome was analyzed.

**Table 2:** Demographics, Clinical, and Clinical Outcome Variables of Subjects Hospitalized with COVID-19 Based on their Use of an Anticoagulant Prescribed as an Outpatient and Continued During Hospitalization or Started New in the Hospital.

| Variable | Sample<br>(N=1716)<br>% | Home Tx AC<br>Continued<br>(N=171)<br>% | Tx AC started<br>without<br>etiology<br>(N=78)<br>% | Tx AC started<br>for new VTE<br>(N=99)<br>% | Tx AC started<br>for new Atrial<br>Fibrillation<br>(N=24)<br>% | VTE<br>Prophylaxis<br>(N=1298)<br>% | No Tx AC or<br>VTE<br>Prophylaxis<br>(N=46)<br>% |
| --- | --- | --- | --- | --- | --- | --- | --- |
| **Demographics** |  |  |  |  |  |  |  |
| Age<sup>a</sup> |  |  |  |  |  |  |  |
| 18-44 | 22.1 | 5.3 | 16.7 | 18.2 | 8.3 | 25.0 | 26.1 |
| 45-59 | 29.6 | 11.1 | 28.2 | 33.3 | 0.0 | 33.0 | 13.0 |
| 60-69 | 21.6 | 22.8 | 25.6 | 25.3 | 33.3 | 20.8 | 17.4 |
| 70-79 | 14.1 | 26.9 | 17.9 | 18.2 | 54.2 | 10.9 | 21.7 |
| >80 | 12.6 | 33.9 | 11.5 | 5.1 | 4.2 | 10.3 | 21.7 |
| Male<sup>b</sup> | 54.5 | 56.1 | 59.0 | 74.7 | 58.3 | 52.7 | 45.7 |
| Race<sup>a</sup> |  |  |  |  |  |  |  |
| White | 35.6 | 63.2 | 32.1 | 23.2 | 45.8 | 32.4 | 52.2 |
| Black | 16.8 | 19.9 | 14.1 | 28.3 | 8.3 | 16.2 | 8.7 |
| Hispanic | 38.6 | 11.7 | 46.2 | 38.4 | 37.5 | 42.0 | 32.6 |
| Other | 8.9 | 5.3 | 7.7 | 10.1 | 8.3 | 9.5 | 6.5 |
| BMI |  |  |  |  |  |  |  |
| Missing weight | 5.7 | 6.4 | 0.0 | 1.0 | 0.0 | 6.2 | 8.7 |
| 18.5-24.9 | 21.4 | 22.2 | 21.8 | 22.2 | 29.2 | 20.9 | 26.1 |
| 25.0-29.9 | 28.3 | 29.8 | 25.6 | 33.3 | 29.2 | 28.1 | 21.7 |
| 30.0-39.9 | 33.0 | 31.6 | 28.2 | 33.3 | 33.3 | 33.3 | 37.0 |
| >40 | 11.7 | 9.9 | 24.4 | 10.1 | 8.3 | 11.5 | 6.5 |
| Charlson score<sup>a</sup> |  |  |  |  |  |  |  |
| 0 | 13.9 | 1.2 | 7.7 | 8.1 | 4.2 | 16.3 | 21.7 |
| 1 | 16.4 | 3.5 | 5.1 | 19.2 | 0.0 | 19.2 | 8.7 |
| 2-3 | 26.1 | 7.6 | 32.1 | 29.3 | 29.2 | 28.5 | 8.7 |
| 4-5 | 16.4 | 21.1 | 21.8 | 18.2 | 20.8 | 15.2 | 17.4 |
| 6-7 | 11.2 | 21.1 | 14.1 | 18.2 | 29.2 | 8.7 | 17.4 |
| >8 | 16.0 | 45.6 | 19.2 | 7.1 | 16.7 | 12.2 | 26.1 |
| **Clinical characteristics** |  |  |  |  |  |  |  |
| Glucose on admission<sup>b,c</sup> |  |  |  |  |  |  |  |
| <81 | 2.3 | 4.1 | 3.8 | 0.0 | 4.2 | 2.2 | 0.0 |
| 81-126 | 51.0 | 53.8 | 32.1 | 40.4 | 29.2 | 52.5 | 63.0 |
| 127-180 | 27.7 | 28.7 | 37.2 | 33.3 | 54.2 | 26.2 | 26.1 |
| 181-350 | 15.8 | 12.9 | 21.8 | 20.2 | 8.3 | 15.9 | 6.5 |
| >351 | 3.1 | 0.6 | 5.1 | 6.1 | 4.2 | 3.1 | 4.3 |
| Antiplatelet agent | 16.5 | 22.8 | 19.2 | 16.2 | 12.5 | 15.6 | 17.4 |
| Hypertension<sup>a</sup> | 58.6 | 84.2 | 61.5 | 65.7 | 83.3 | 53.9 | 63.0 |
| Diabetes<sup>a</sup> | 34.7 | 44.4 | 56.4 | 36.4 | 25.0 | 32.6 | 21.7 |
| Asthma | 10.7 | 12.3 | 11.5 | 6.1 | 16.7 | 11.0 | 2.2 |
| Chronic obstructive pulmonary disease<sup>a</sup> | 10.7 | 24.6 | 16.7 | 9.1 | 20.8 | 8.5 | 10.9 |
| Kidney disease<sup>a</sup> | 19.3 | 41.5 | 21.8 | 23.2 | 29.2 | 15.6 | 23.9 |
| Congestive heart failure<sup>a</sup> | 15.4 | 53.8 | 17.9 | 18.2 | 25.0 | 9.6 | 23.9 |
| History of stroke<sup>a</sup> | 5.9 | 19.9 | 5.1 | 5.1 | 12.5 | 4.1 | 6.5 |
| Peripheral artery disease<sup>a</sup> | 5.8 | 14.0 | 7.7 | 5.1 | 8.3 | 4.7 | 4.3 |
| Coronary artery disease<sup>a</sup> | 5.1 | 13.5 | 7.7 | 3.0 | 4.2 | 3.8 | 13.0 |
| Malignancy (solid /heme) | 10.10 | 19.30 | 11.50 | 10.10 | 8.30 | 8.50 | 21.70 |
| Immunodeficiency | 1.3 | 1.8 | 0.0 | 1.0 | 8.4 | 1.3 | 2.2 |
| **Outcomes** |  |  |  |  |  |  |  |
| Overall Death<sup>a</sup> | 7.3 | 12.3 | 37.2 | 13.1 | 16.7 | 4.1 | 10.9 |
| Critical illness<sup>a</sup> | 32.3 | 36.3 | 85.9 | 71.7 | 75.0 | 25.1 | 23.9 |
| Mechanical Ventilation<sup>a</sup> | 14.8 | 12.9 | 60.3 | 51.5 | 37.5 | 9.4 | 6.5 |
| Death in critical illness<sup>a,d</sup> | 19.3 | 24.2 | 43.2 | 23.9 | 22.2 | 12.6 | 45.5 |
| No AC= did not receive anticoagulation<br>Tx=Therapeutic<br>AC=Anticoagulation<br>VTE= Venous thromboembolism<br>a. $P<0.001$<br>b. $P<0.01$<br>c. 2 non-diabetic subjects did not have admission glucose levels (their closest glucose level to admission was used for analysis – 1 from 2 months before and 1 at 1 year before admission)<br>d. Group included only critically ill subjects (Total: N=555; Home AC: N=62; Tx AC started without etiology: N=67; Tx AC started for new VTE: N=71; Tx AC started for new atrial fibrillation: N=18; VTE prophylaxis: N=326; No AC: N= 11) |  |  |  |  |  |  |  |

We compared subjects’ prescribed therapeutic anticoagulants (either continued from home or started in the hospital) to prophylactic anticoagulants on four clinical outcomes: overall death (primary outcome), critical illness (as defined as scores ≥5 on the WHO COVID-19 eight-point ordinal scale), need for mechanical ventilation, and death in critical illness (subjects who first had critical illness before dying; WHO score 5-7) (secondary outcomes).

### Measurement

The percentage of overall deaths was calculated as the number of subjects who died divided by the *total* number of discharges and deaths. The percentage of subjects with critical illness, mechanical ventilation, and death among subjects who first had critical illness was calculated as the number of subjects with each outcome divided by the number of critically ill subjects who were discharged or died. Subjects who died before first detecting critical illness were excluded in the analysis of critical illness status, mechanical ventilation, or death in critical illness because confounding variables may have prevented transfer to ICU (do-not-resuscitate status or treatment refusal).

### Analysis

We calculated the Charlson comorbidity index for each admission (21,22). We used the Chi-square test to determine the significance of differences in baseline demographic and clinical characteristics and clinical outcomes between the six anticoagulation therapy groups. A p-value of <0.01 was used as the threshold for statistical significance in these bivariate analyses to adjust for multiple comparisons. Poisson regression was used to estimate associations between subject attributes and exposures and the likelihood of death, as the incidence rate ratio is a closer approximation to relative risk given the observed 7.3% overall death rate in this cohort (23). Multiple logistic regression was used to determine associations between the six anticoagulation therapy groups with the likelihood of critical illness, need for mechanical ventilation and death amongst those with critical illness. Prophylactic anticoagulation was used as the reference group in regression models as this is the standard of care in our hospital. Regression models were adjusted for age, sex, race and ethnicity, body mass index (BMI), Charlson score, glucose on admission, and use of antiplatelet agents. Age, BMI, Charlson score, and blood glucose were categorized for ease of interpretation of odds ratios (24,25). Sensitivity analyses were performed for all four regression models controlling for pre-existing hypertension, diabetes, asthma, chronic obstructive pulmonary disease, kidney disease, congestive heart failure, stroke, peripheral vascular disease, coronary artery disease, malignancy, or immunodeficiency in place of the Charlson score. All statistical analyses were performed using SPSS® Version 26 (Armonk, NY) and Stata Version 15.1 (College Station, TX).

## RESULTS

We identified 1,716 adults hospitalized with COVID-19 of whom 372 were on therapeutic anticoagulation during their admission. Manual chart review for each of these subjects confirmed their COVID-19 diagnosis and assigned each subject to one of the six anticoagulation groups. Therapeutic anticoagulation had been prescribed prior to hospitalization and was subsequently continued during hospitalization for 171 subjects (10.0%). Therapeutic anticoagulation was initiated de novo during hospitalization for 201 subjects (11.7%). Prophylactic anticoagulation was given during hospitalization for 1,298 (75.6%). 46 subjects (2.7%) did not receive therapeutic or prophylactic anticoagulation. Of the 201 subjects newly started on therapeutic anticoagulation while hospitalized, 99 (5.8 %) were treated for new VTE, 24 (1.4 %) for new atrial fibrillation, and 78 (4.5%) for COVID-19 disease without other indications. Of the 78 subjects who received therapeutic anticoagulation for COVID-19, total duration of anticoagulation was a median 4 days [Interquartile range (IQR) 2-7 days].

In bivariate analyses (Table 2), those subjects continued on prior anticoagulation were more likely to be older, have higher Charlson scores, and have more frequent comorbid conditions compared to those beginning prophylactic anticoagulation while hospitalized. Subjects started on therapeutic anticoagulation compared to those on prophylactic anticoagulation were more likely to have higher Charlson score and more frequent comorbid conditions. Table 2 also shows that subjects on anticoagulation therapy (both continued outpatient and new inpatient) had worse primary outcomes of overall death and secondary outcomes of critical illness, mechanical ventilation, and death in critically ill compared to those on prophylactic doses. One hundred twenty-five subjects in our health system died due to COVID-19 disease (7.3%). Eighteen deaths occurred without critical illness status (WHO score 3-4) and 107 deaths occurred in those who first had critical illness (WHO score 5-7). The mortality rate was 19.3% in critically ill subjects.

After adjusting for covariates, there was no difference in death, critical illness, mechanical ventilation, or death in critical illness among subjects continued on therapeutic anticoagulation when compared to subjects who received prophylactic anticoagulation (Table 3). These results were unchanged in the sensitivity analysis. Of note, subjects who started on new therapeutic anticoagulation in the hospital had the worse clinical outcomes (Table 3). Of the subjects who received therapeutic anticoagulation for COVID-19 infection, 85.9% had a critical illness status with an increased risk of overall death (OR 5.93; 95% CI 3.71-9.47), critical illness (OR 14.51; 95% CI 7.43-28.31), mechanical ventilation (OR 11.22; 95% CI 6.67-18.86), and death in critical illness (OR 5.51; 95% CI 2.80 −10.87). Therapeutic anticoagulation started for new VTE was associated with increased risk of overall death (OR 3.13; 95% CI 1.66-5.93), critical illness (OR 6.75; 95% CI 4.19-10.87) and needing mechanical ventilation (OR 9.03; 95% CI 5.66-14.41). Therapeutic anticoagulation started for new atrial fibrillation had increased risk of secondary outcomes of critical illness (OR 6.47; 95% CI 2.44-17.17) and needing mechanical ventilation (OR 3.97; 95% CI 1.61-9.81). Increased age, male sex, BMI and increased Charlson score was associated with worse outcomes (Table 3).

**Table 3:**
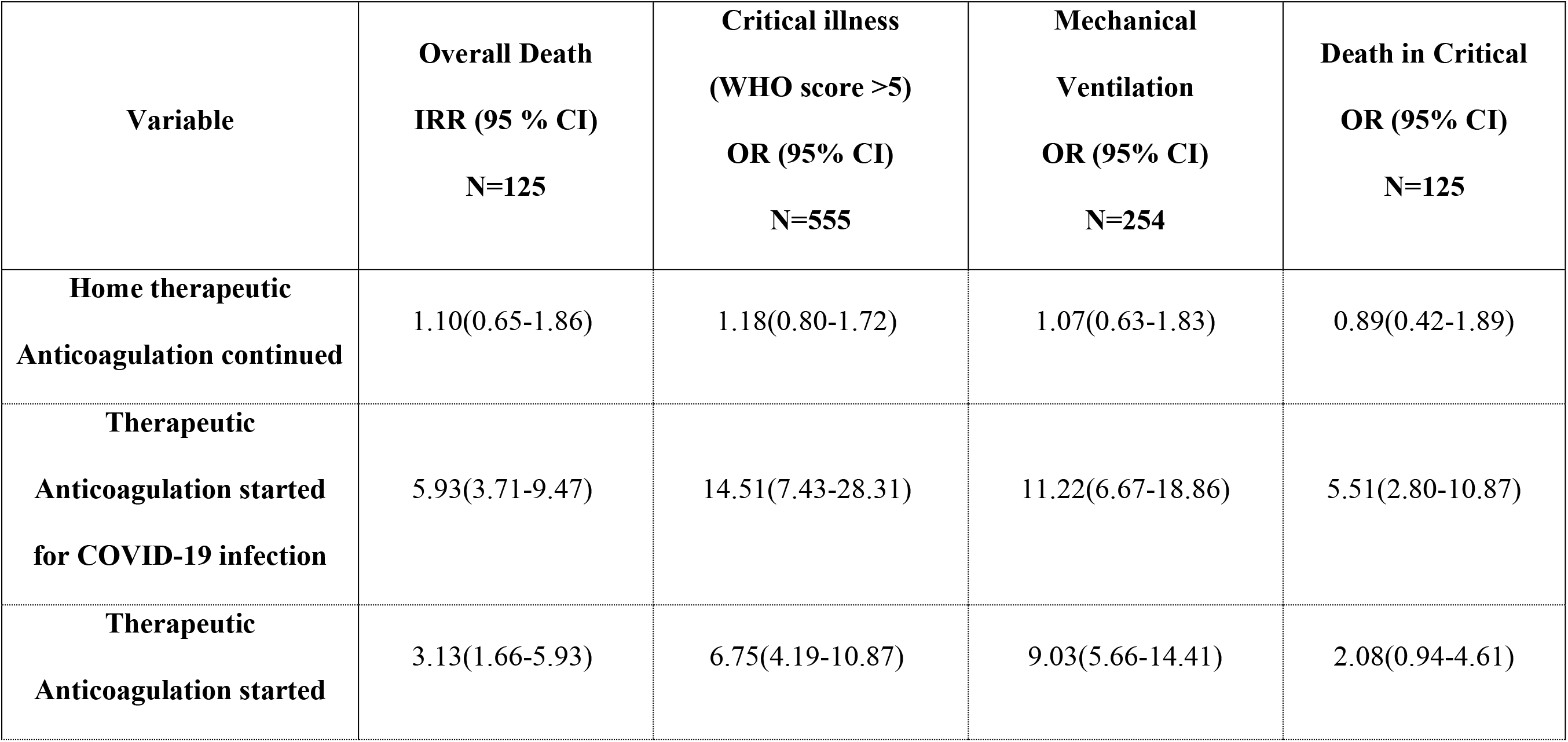
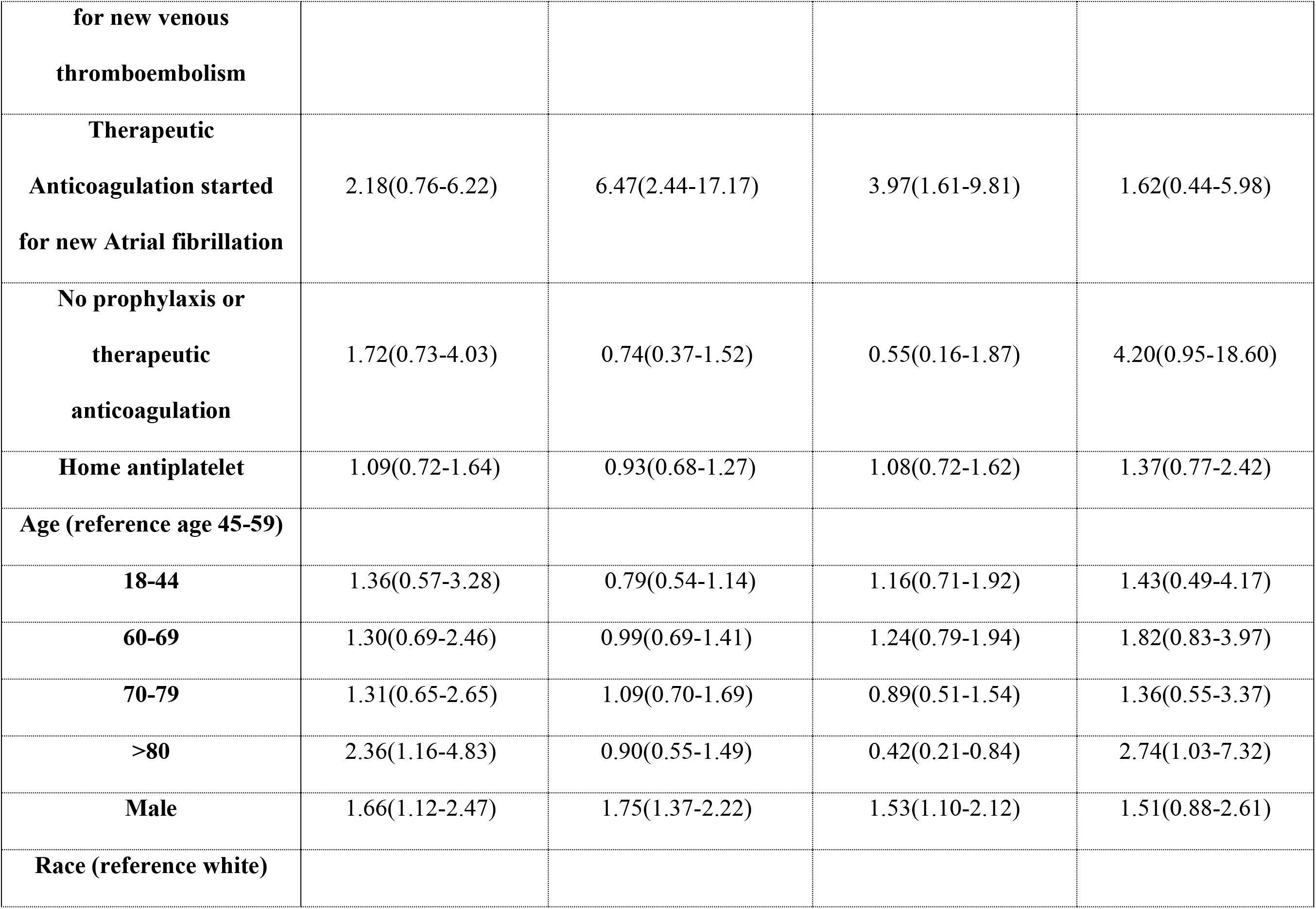
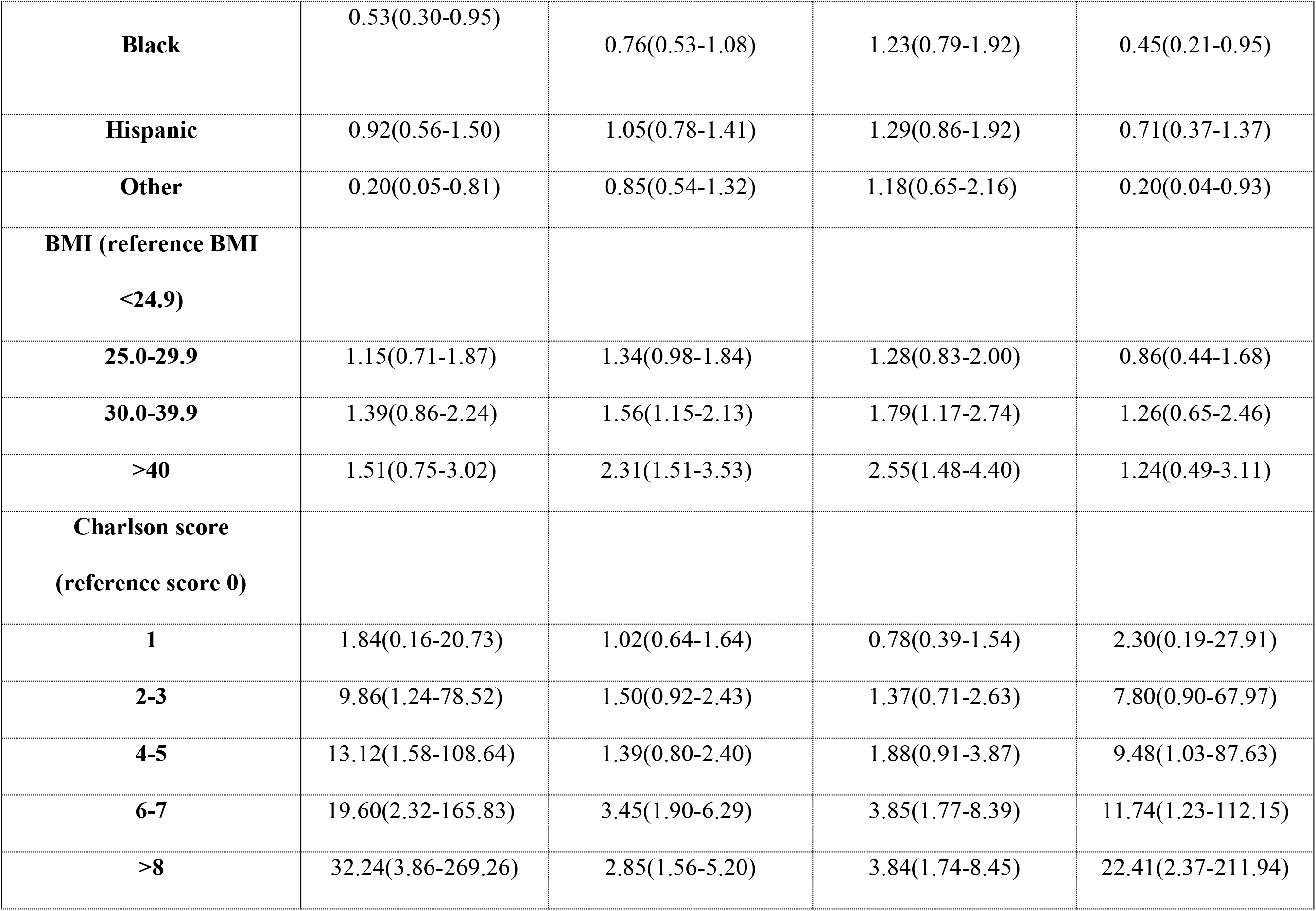
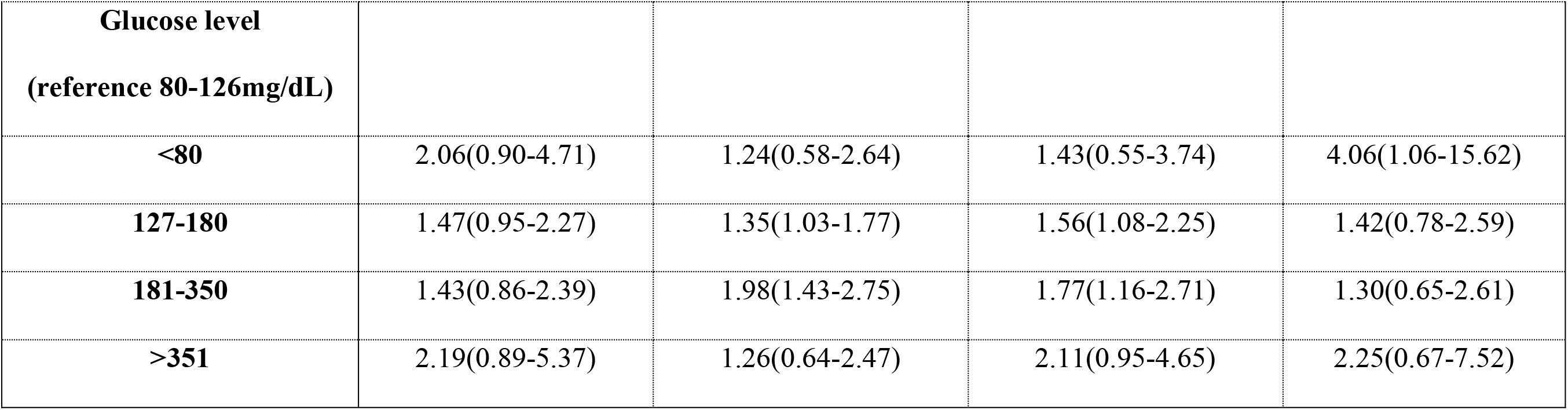
Poisson and Multiple Logistic Regression Analyses of 1716 Subjects Hospitalized with COVID-19 the Incidence Rate Ratio [IRR; (95% CI)] or the Odds Ratios [OR; 95% Confidence Interval (CI)] of Subject Anticoagulation Status with Overall Death, Critical Illness, Mechanical Ventilation, and Death among Critical Subjects.

## DISCUSSION

Subjects hospitalized with COVID-19 who were prescribed therapeutic anticoagulation prior to hospitalization and who were continued on therapeutic anticoagulation while hospitalized had no difference in primary outcomes of overall death compared to subjects who started prophylactic anticoagulation while hospitalized; there was also no difference between these groups for secondary outcomes of critical illness, mechanical ventilation, or death in critical illness. Subjects who were started on anticoagulation for COVID-19 without evidence of thrombosis, new VTE, or new atrial fibrillation had worse outcomes compared to subjects who were on prophylactic anticoagulation.

Our study findings add to a growing body of observational studies evaluating the effects of therapeutic anticoagulation in patients with COVID-19. In the study by Parajpe et al., patients on mechanical ventilation were less likely to die if they were on therapeutic anticoagulation (29.1% vs. 62.7%) (14). In two other observational study by Tang et al. and Ayerbe et al., patients with COVID-19 had a mortality benefit if they received low molecular weight heparin (LMWH) or heparin, respectively (15,26). However, neither of these two studies specified whether the dose of anticoagulant was therapeutic or prophylactic. Two other studies limited by low numbers reported lower mortality in patients receiving therapeutic anticoagulants (27,28). In contrast, a study of nursing home patients suggested that continuation of oral therapeutic anticoagulation in patients with COVID-19 infection did not have a mortality benefit (29). Additionally, another observation study posted on medrxiv.org (that has not yet undergone peer review) reported a higher mortality rate among patients with COVID-19 who received t empiric therapeutic anticoagulation with other indications (30). Based on current evidence, we do not believe firm recommendations for or against empiric anticoagulation in patients with COVID-19 can be made. In our hospitals, we do not routinely use therapeutic anticoagulation for patients with COVID-19 infection without other indications (31,32).

Our study had other significant findings. First, of the subjects who were not already on therapeutic anticoagulation, 6.4% of subjects with COVID-19 were anticoagulated for a newly diagnosed VTE similar to other studies on the incidence of VTE in COVID-19 (4,33). Second, our study confirms previously identified risk factors for worse outcomes in COVID-19 including sex, BMI, blood glucose levels, and comorbid conditions (20,34,35). However, the overall mortality rate (7.3%) of subjects in our study was lower than that reported in other studies. Recent studies out of the New York City area and Louisiana showed inpatient mortality rates between 21-24% (35,36). Our mortality rate may be lower because our health system never became overwhelmed, patients were cared for by a multi-disciplinary team that rounded twice daily, and clinicians in our ICUs treated ventilated patients with low volume, high peep strategies as well as prone positioning (37,38).

In our cohort, dexamethasone was not widely used during the study period. It seems plausible that routine prophylaxis against or treatment of endotheliïtis with glucocorticosteroids may result in reduced incidence of thrombosis with reductions in overall mortality, as was reported in the RECOVERY study (39,40). However, the optimal timing of administration of dexamethasone during COVID-19 illness is not clear as glucocorticosteroids may also suppress innate and adaptive immune response, which may result in higher viral load and greater excursion of blood glucose values, leading to worse outcomes in some patients (41,42).

Our study has several limitations. First, our-cohort definition and clinical data relied on the accuracy of ICD-10 coding for disease diagnoses, although we did manually review 372 (22%) charts which confirmed COVID-19 infection and assessed timing and indication for therapeutic anticoagulation. Second, our study was observational, and despite attempting to control for confounding clinical variables, there may have been unidentified confounding factors. Third, we did not evaluate outcomes by individual anticoagulant drug. It is possible that the clinical effects of DOACs and LMWH differ in patients with COVID-19 (27,43). Finally, we did not control for the number of days subjects received new therapeutic anticoagulation because we were unable to determine the timing of starting these medications within the course of COVID-19 infection.

In conclusion, continuation of home therapeutic anticoagulation was associated with neither benefit nor harm in subjects hospitalized with COVID-19. Subjects with COVID-19 who had not previously been anticoagulated and who were empirically prescribed therapeutic anticoagulation during their hospitalization had worse outcomes. Our findings do not support routine use of therapeutic anticoagulation for patients with COVID-19 infection in the absence of other indications (i.e., atrial fibrillation or venous thromboembolism) until randomized controlled trials provide further evidence.

## Data Availability

Data is not currently publicly available as it contains patient health information and is not to be shared with authors not involved in the study per the IRB.

## Acknowledgements

We would like to acknowledge Dr. Douglas Vaughan for his support of this project and Dr. Brady Stein for editorial feedback on the manuscript. We thank the countless clinicians, environmental and food services personnel, and other staff at Northwestern Medicine for their dedication to patient care during the COVID-19 pandemic.

